# PRACTICES OF OCCUPATIONAL HEALTH AND SAFETY AND EXPERIENCE OF OCCUPATIONAL HAZARDS AMONG SOLID WASTE MANAGERS IN PORT HARCOURT METROPOLIS OF RIVERS STATE

**DOI:** 10.1101/2023.02.17.23285698

**Authors:** Stanley Nyemahame Amadi, Emmanuel Clement, Emmanuel Tamunobelema Pelebo, Uchechukwu Ifeanyichukwu Apugo

## Abstract

**Background:** Waste generation is a daily occurrence and requires a proper system of collection, management, and disposal. This function lies on municipal solid waste managers (MSWM) who use manual methods of waste collection and management in the conduct of their activities. This exposes them to various forms of occupational health risks.

**Aim:** The study investigated the occupational risk exposure of waste managers in Port Harcourt metropolis.

**Materials and Method:** This descriptive cross-sectional survey collected data from solid waste collectors or managers who have worked for over a year in the Rivers state, using a researcher-administered semi-structured questionnaire. The data was analysed using SPPS version 23.

**Results:** A total of 317 were recruited, with the majority as males (68.5%), aged between 30 – 39 years (56.5%), married (55.5%), permanent staff (54.3%), educated to the tertiary level (31.2%), have over 5 years of experience (52.1%) and earn between 30,000 – 39,000 Naira monthly (44.8%). Also, 87.1% of the respondents had good practice of occupational health and safety, while 61.5% and 32% of the waste managers have been exposed to work-related accidents and diseases respectively in course of carrying out their job within the last 12 months. Cuts (30.0%), puncture wounds (20.5%), and road accidents (20.2%) as the most experienced hazards by the workers, while allergies (23.2%) and rash/other skin diseases (22.1%) were the most experienced type of illness.

**Conclusion:** The high prevalence of occupational hazards can be attributed to poor working conditions and lack of adequate safety gear. Hence, there is a need for increased governmental budgetary allocation for the provision safety gear and training

## INTRODUCTION

indiscriminate waste disposal methods and inadequate waste collection has continuously posed as the most intractable problem of waste management (WM) in Nigeria, especially in the Niger Delta Region.^1,2^ According to the United Nations Environment Programme (UNEP), most cities in the world produce an average of 1.9 billion tons of Solid Waste (SW) yearly, with those in Sub-Saharan Africa (SSA) producing an estimated 62 million tons per year.^3^ Hoornweg and Bhada-Tata^4^ added that cities throughout the world produced 1.3 billion tons of SW in 2012, with that figure anticipated to rise to 2.2 billion tons by 2025. This is plainly too much for local waste management authorities to handle, due to the employment of inadequate labour-intensive (manual) procedures that forms the modus operandi in many developing countries. Though this manual method of SW collection and management has some advantages in terms of employment, it has also been known to cause occupational injuries and diseases such as acute diarrhoea, cholera, dysentery, injuries from sharp objects, malaria, respiratory disease, typhoid fever, strains from lifting, and contact with pathogens for those who collect anaerobic waste.^5^ It is therefore unsurprising that this hazardous job has a death rate of about 90 per 100,000 garbage collectors globally.^6^

Solid Waste Collectors (SWCs) are frequently engaged in risky working circumstances with poor pay and minimal social protection. Hence, Madian and El-Wahed^6^ stated that the occupation is ranked as the seventh most dangerous occupation in the world. Some of the contributors to this outcome include the fact that the job requires a lot of physical efforts, such as hand lifting and carrying of heavy bins, exposure to sharp metals, and involves operating on a truck that goes through traffic all year.^7^ Such individuals are also exposed to an array of pathogenic substances (bacteria, fungi, viruses, parasites, and cysts), toxic substances, waste-related and decomposition-related chemicals, as well as vehicle exhaust fumes, noise, extreme temperatures, and UV radiation, in the course of carrying out the jobs.^8^ This is more prominent in low- and middle-income countries (LMICs) like Nigeria., as the World Health Organization (WHO) reported that SWCs in developing countries are three times more likely to develop allergic respiratory disease, one and a half times more likely to develop non-allergic respiratory disease, three times more likely to develop chronic bronchitis, and six times more likely to develop infectious diseases.^6,9^ Hence, this study was set at determining the prevalence of the experience of occupational hazards among the solid waste collectors or managers in Port Harcourt Metropolis and their practice of Occupational Health and Safety (OHS) in the process of carrying out their jobs.

## METHODOLOGY

### Study Design and setting

This descriptive cross-sectional survey was conducted in Port Harcourt metropolis, which is located in Rivers State between Latitude 4°45 ’N and Latitude 4°55 ’N, and Longitude 6°55 ’E and Longitude 7°05 ’E. The metropolis is made up of two (2) local government areas (LGAs); Port Harcourt and Obio/Akpor, which are the state’s two most populous urban LGAs. The population boom in the metropolis has resulted in a densely populated area with poor design, with dwellings erected closer together than ever before. As a result, about 80% of solid trash created comes from household sources.

### Study Population

The target population for this study consisted of solid waste collectors or managers (irrespective of age and gender), employed by the Rivers State Waste Management Authority (RIWAMA).

### Sample Size Determination and Sampling Techniques

The needed minimum sample size (n) of 317 was calculated using the Cochran equations with p as the prevalence of occupational hazards among waste managers (76%),^10^ 5% margin of error, 95% confidence interval, and considering a 10% non-response rate. The respondents were recruited using a multi-staged sampling method

### Study Instruments

This study used a semi-structured questionnaire that gathered information on the socio-demographic and work-related characteristics of the SWCs, their experience of work-related hazards or injury, and the Practice OHS guidelines while carrying out their routine tasks.

### Statistical Analysis

Data from completed surveys was analysed using the Statistical Package for Social Science (SPSS) version 23. The results were derived using descriptive statistics and provided as a frequency count and percentage for socio-demographic information, work-related factors, and the presence and kind of risks to which waste managers are exposed.

### Ethical Consideration

he University of Port Harcourt’s Research and Ethics Committee approved the conduct of this study while permission was granted by RIWAMA. The respondents also gave written informed consent.

## RESULTS

### Socio-demographic and work-related characteristics

The majority of waste managers recruited into this study were aged between 30 – 39 years (56.5%), males (68.5%), married (55.5%), educated to the tertiary level (31.2%), employed as permanent workers (54.3%), worked for over 5 years (52.1%), received monthly income of about 30,000 – 39,000 Naira (44.8%), involved in sorting wastes (62.1%), work for 4 – 5 days weekly (55.8%), and spend 7 – 10 hours daily on the solid waste vehicles (37.9%).

**Table 1:** Socio-demographic and work-related characteristics of waste managers in Port Harcourt

| **Variables (N = 317)** | **Frequency** | **Percentage** |
| --- | --- | --- |
| **Age category** |  |  |
| ≤29 years | 46 | 14.5 |
| 30 – 39 years | 179 | 56.5 |
| 40 – 49 years | 79 | 24.9 |
| ≥50 years | 13 | 4.1 |
| **Sex** |  |  |
| Male | 217 | 68.5 |
| Female | 100 | 31.5 |
| **Marital status** |  |  |
| Single | 117 | 36.9 |
| Married | 176 | 55.5 |
| Divorced | 10 | 3.2 |
| Widowed | 14 | 4.4 |
| **Educational status** |  |  |
| None | 17 | 5.4 |
| Primary | 77 | 24.3 |
| Secondary | 99 | 31.2 |
| Tertiary | 124 | 39.1 |
| **Nature of employment** |  |  |
| Permanent | 172 | 54.3 |
| Temporary | 145 | 45.7 |
| **Years of working experience** |  |  |
| 1 year | 7 | 2.2 |
| 2 – 3 years | 26 | 8.2 |
| 4 – 5 years | 119 | 37.5 |
| >5 years | 165 | 52.1 |
| **Monthly Income (NGN)** |  |  |
| <20,000 | 41 | 12.9 |
| >20,000 – 29,000 | 134 | 42.3 |
| 30,000 – 39,000 | 142 | 44.8 |
| **Involvement in the sorting of waste** | 197 | 62.1 |
| **Number of days at work in a week** |  |  |
| 2 – 3 days | 20 | 6.3 |
| 4 – 5 days | 177 | 55.8 |
| 6 – 7 days | 120 | 37.9 |
| **Number of hours spend in a solid waste vehicle** |  |  |
| 1 – 3 hours | 49 | 15.5 |
| 4 – 6 hours | 77 | 24.3 |
| 7 – 10 hours | 120 | 37.9 |
| >10 hours | 71 | 22.4 |

### Practice of Occupational Health and Safety

Analysis of the result of the practice of OHS as presented in figure 1 showed that 87.1% of the respondents had a good level of practices of OHS while 12.9% had a poor level of OHS practices. the result of the occurrence of work-related injuries as presented in figure 2 above revealed that 195 (61.5%) of the respondents had experienced work-related injuries/accidents within the last 12 months, with cuts (30.0%) puncture wounds (20.5%) and road accidents (20.2%) as the most experienced hazards.

**Table 1:**
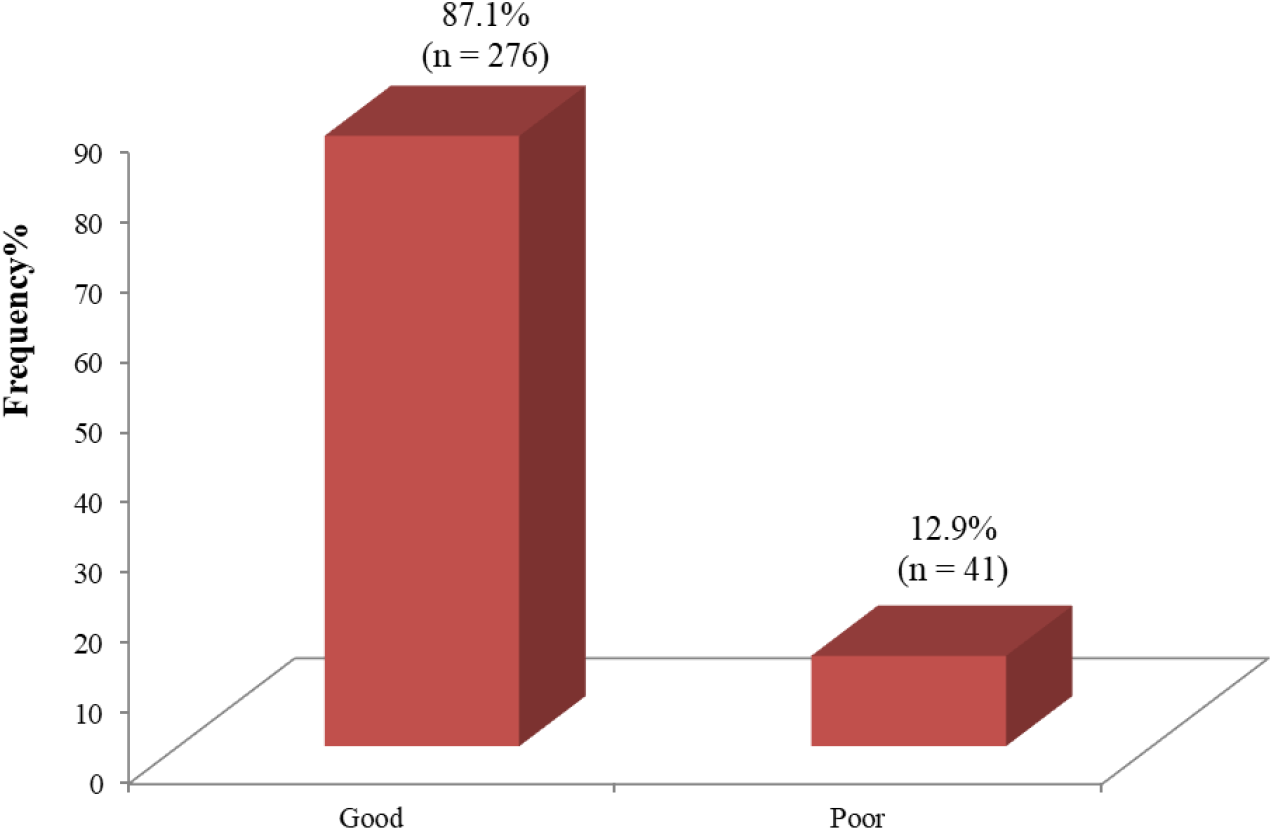
Practice of occupational health and safety by waste managers in Port Harcourt

**Figure 2:**
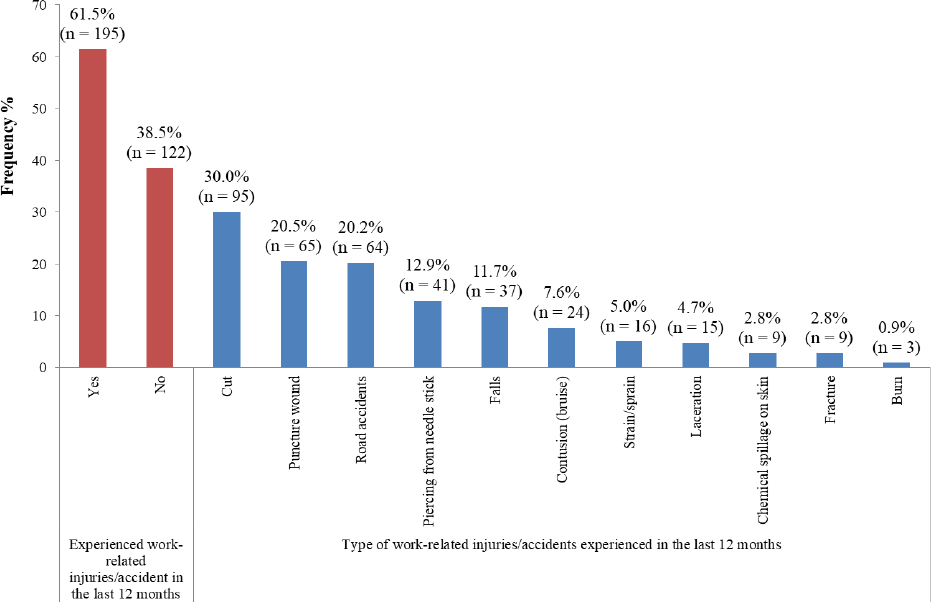
Experience of work-related injuries/accidents and the type of injuries/accidents

### Experience of OHS

Figure 3 above presents the rate of hospitalization due to work-related hazards and illness. According to the result, 32.2% of the respondents reported that they have even been hospitalized due to work-related hazards, with allergies (23.2%) and rash/other skin diseases (22.1%) identified as the most experienced type of illness

**Figure 3:**
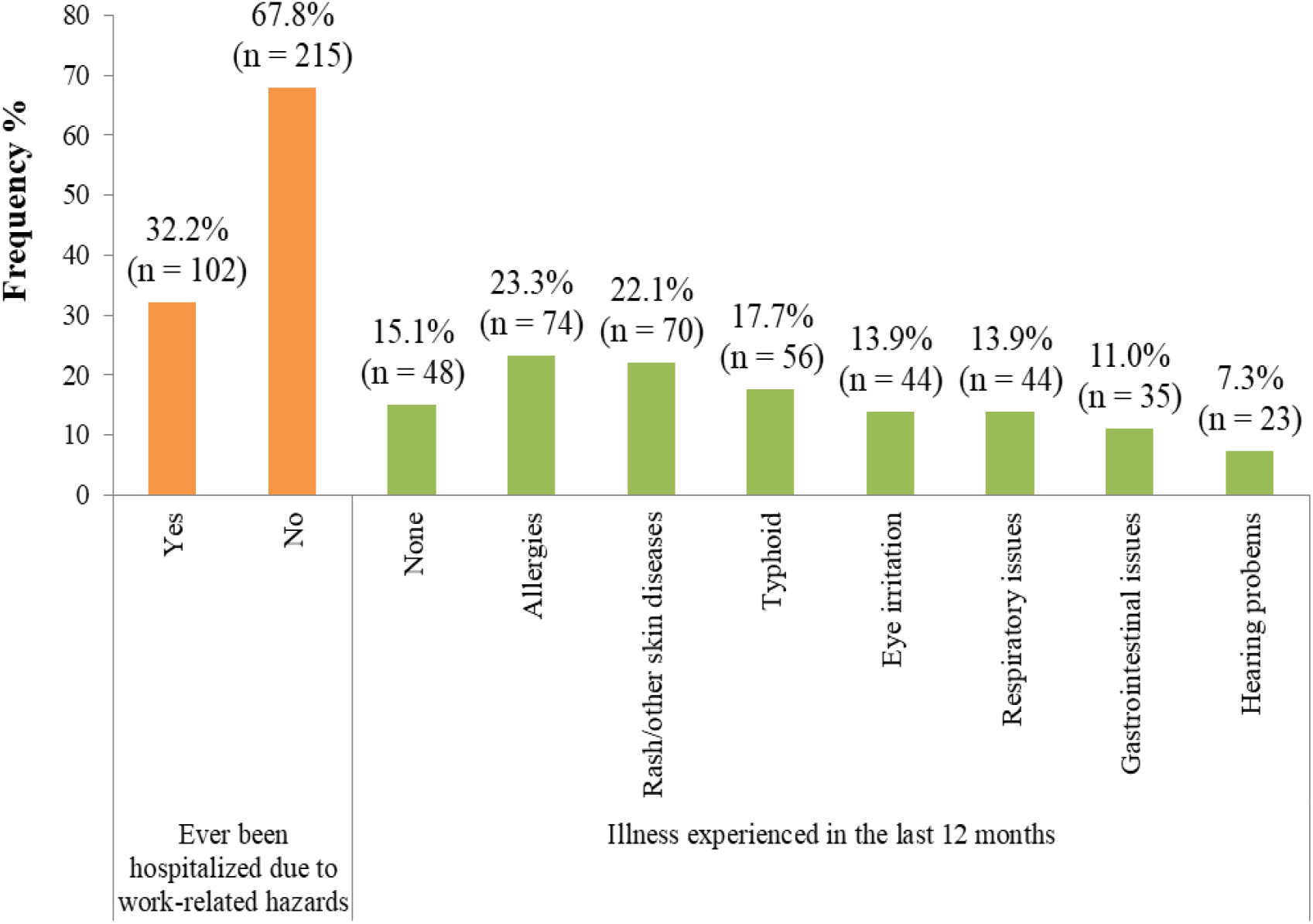
Rate of hospitalization due to work-related hazards and illness ever been experienced

## Discussion of Result

Poor working conditions and a lack of protective equipment lead to higher accident rates among waste managers in most developing nations, particularly in SSA and Asia.^20^ According to Aweng and Fatt,^21^ the process of managing waste entails a wide variety of activities, with various types of dangers present at every stage of the process, from the point of collection at households through transportation and recycling or disposal locations.^6^ Though PPEs may not completely avoid exposure to various types of accidents, their frequent use by waste handlers would greatly lower the likelihood of exposure to hazardous compounds contained in the waste stream.^11,12^ The majority of waste managers in this study had good OHS practices (87.1%), while the total usage of PPE was 90.9%, with the frequency of use being occasionally (42.0%) and hand gloves being the most commonly used PPE. This is higher in comparison with the results reported in the studies of Kretchy^13^ in Ghana and Wahab and Ogunlolo^14^ in Ekiti state, Nigeria, who found 72.6% and 60% excellent safety procedures, respectively. Also, this finding outperformed Bogale and Tefara^15^ and Miwano et al^.10^ who found that only 22%, 30%, and 24.06% of SWC had effective safety procedures, respectively. Sawyerr et al.^16^ found that the majority (91.7%) of municipal SWCs in Ilorin, Nigeria, did not utilize PPE, while Madian and El-Wahed^6^ reported that 65% of the SWC in their study used PPE on the job, with safety boots being the most commonly used PPE (73.8%). Diwe et al.^17^ did report that the overall PPE use in their study was 66.7%, with 56.1% using it always, while Bogale et al.^15^ found that just 43.6% of SWC used PPE, with only a fifth reporting that they did not use it on a regular basis. This high level of good practice and PPE usage among respondents might be attributed to governmental enforcement, effective worker education or pre-job training, and careful monitoring to ensure that particular laid-down criteria are followed.^10,13,14^ The limited use of PPE, on the other hand, can be attributed to a lack of assistance from the necessary authorities in terms of providing particular kits.^10^

In the current research, 61.5% of the SWCs said they had been exposed to work-related incidents such as cuts, punctures, and traffic accidents in the past 12 months while doing their duties. This is higher in comparison with the findings of Alshebli et al.^22^ where 56% of SWC were highly exposed to occupational accidents, while the studies of Marahatta et al.^23^ and Madian and El-Wahed^6^ reported higher exposure of SWCs to work-related accidents (80% and 76% respectively), with needle stick injury, falls, fissure feet and contusions, puncture wounds and lacerations, fracture, animal bites, and chemical injuries reported as the most frequently occurring. Similarly, Eskezia et al.^24^ also reported the occurrence of injuries such as cuts from sharp equipment on their hands or legs, and hand fractures as a result of falling among the SWCs in their study. The occurrence of these types of injuries could be associated with a lack of waste segregation and appropriate disposal mechanisms by households and even healthcare facilities. Also, most dumpsites are located along the roads and this exposes the workers to road traffic accidents.

In the area of work-related illnesses and diseases, 32% of the SWCs stated that they had been hospitalized in the previous year due to work-related diseases such as allergies, rashes, typhoid, eye irritation, and other respiratory difficulties. In contrast to this, an Indian study by Jayakrishnan, Jeeja, and Bhaskar^25^ found that only 13.1% of SWCs in their study had been hospitalized for a work-related illness, while Madian and El-Wahed^6^ reported that almost half of the managers in their study had previously been hospitalized due to a work-related illness, with gastrointestinal issues (worm infection), eye issues (redness), skin issues (itching, nail infection, and scabies), respiratory issues (cough and dyspnea), and musculoskeletal discomfort (neck and low back pain) among them. Purushottam et al.^26^ reported that 89% of waste managers in Bombay city had eye problems (burning sensation, watering redness, and itching), while Abou-Elwafa et al.^27^ and Garrido et al.^28^ found that 90% and 61% of waste managers in Germany and Egypt, respectively, had musculoskeletal complaints. Cough, cholera, asthmatic attacks, trouble breathing, skin disorders, eye irritation, dysentery, and typhoid fever were reported by waste managers in Lagos and Ibadan.^12^

In accordance with this report, Madian and El-Wahed^6^ reported that the amount of time SWCs spend executing their normal operations increases their sensitivity to various ailments and the need for hospitalization, while Malta-Vacas et al.^29^ reported that working with waste for longer periods of time causes serious GI problems due to their exposure to a variety of diseases, vectors, and direct skin contact with materials containing flora spores, bacteria, viruses, and parasitic ova, all of which cause gastrointestinal discomfort. Also, Otitoju^18^ stated that waste management places a significant strain on the musculoskeletal system, particularly the back, while Reddy and Yasobant^19^ and Thakur and Ganguly^11^ stated that this could be as a result of carrying, pulling/pushing of bins and containers, which involves static muscle contraction and increases the risk of musculoskeletal disorders and hospitalisation.

## Conclusion and Recommendation

This study reported a high practice of OHS and usage of PPEs (especially hand gloves), as well as a high rate of occurrence of work-related accidents (cuts, punctures, and traffic accidents) and a lower rate of occurrence of work-related illnesses and diseases (allergies, rashes, typhoid, eye irritation, and other respiratory difficulties) in the last 12 months before the study. This calls for increased governmental budgetary allocation and Non-governmental organisation (NGO) involvement in this sector in the area of the provision of complete and appropriate PPEs meant for the SWCs in the process of carrying out their work. Also, necessary OHS training and retraining tailored specifically for SWCs is required to help enlighten them more on the hazards associated with their work as well as the importance of compliance with PPEs.

## Data Availability

All data produced in the present study are available upon reasonable request to the authors

## Conflicts of interest

The authors hereby declare no conflict of interest.

## Notes

### Competing Interest Statement

The authors have declared no competing interest.

### Funding Statement

This study did not receive any funding

### Author Declarations

Ethics committee/IRB of the University of Port Harcourt gave ethical approval for this work

